# Existence of SARS-CoV-2 RNA on ambient particulate matter samples: A nationwide study in Turkey

**DOI:** 10.1101/2021.01.24.21250391

**Authors:** Özgecan Kayalar, Akif Arı, Gizem Babuççu, Nur Konyalılar, Özlem Doğan, Füsun Can, Ülkü A. Şahin, Eftade O. Gaga, S. Levent Kuzu, Pelin E. Arı, Mustafa Odabası, Yücel Taşdemir, S.Sıddık Cindoruk, Fatma Esen, Egemen Sakın, Burak Çalışkan, Lokman H. Tecer, Merve Fıçıcı, Ahmet Altın, Burcu Onat, Coşkun Ayvaz, Burcu Uzun, Arslan Saral, Tuncay Döğeroğlu, Semra Malkoç, Özlem Ö. Üzmez, Fatma Kunt, Senar Aydın, Melik Kara, Barış Yaman, Güray Doğan, Bihter Olgun, Ebru N. Dokumacı, Gülen Güllü, Elif S. Uzunpınar, Hasan Bayram

## Abstract

Coronavirus disease 2019 (COVID-19) is caused by the SARS-CoV-2 virus and has been affecting the world since the end of 2019. Turkey is severely affected with the first case being reported on March 11^th^ 2020. Ambient particulate matter (PM) samples in various size ranges were collected from 13 sites including urban and urban background locations and hospital gardens in 10 cities across Turkey between the 13^th^ of May and the 14^th^ of June, 2020 to investigate a possible presence of SARS-CoV-2 RNA on ambient PM. A total of 155 daily samples (TSP, *n=80*; PM_2.5_, *n=33*; PM_2.5-10_, *n=23*; PM_10_, *n=19*; and 6 size segregated, *n=48*) were collected using various samplers in each city. The N1 gene and RdRP gene expressions were analyzed for the presence of SARS-CoV-2 as suggested by the Centers for Disease Control and Prevention (CDC). According to RT-PCR and 3D-RT-PCR analysis, dual RdRP and N1 gene positivity were detected in 20 (9.8 %) of the samples. The highest percentage of virus detection on PM samples was from hospital gardens in Tekirdağ, Zonguldak, and İstanbul—especially in PM_2.5_ mode. Samples collected from two urban sites were also positive. Findings of this study have suggested that SARS-CoV-2 may be transported by ambient particles especially at sites close to the infection hot-spots. However, whether this has an impact on the spread of the virus infection remains to be determined.

**Significance Statement:** Although there are several studies reporting the existence of SARS-CoV-2 in indoor aerosols is established, it remains unclear whether the virus is transported by ambient atmospheric particles. The presence of the SARS-CoV-2 RNA in ambient particles collected from characteristic sites within various size ranges was investigated, and positive results were found in urban sites especially around Turkish hospitals. In this context, this study offers a new discussion on the transmission of the virus via ambient particles.

## Introduction

After the first reported cases of unknown pneumonia in Wuhan, China, in December 2019, the World Health Organization (WHO) announced an International Public Health Emergency in January and a pandemic in March 2020. A novel and previously unknown severe acute respiratory syndrome related coronavirus 2 (SARS-CoV-2) was isolated from the epithelial cells of the patients with pneumonia. This was later named Coronavirus Disease 19 (COVID-19) by the WHO in February 2020 (1-3). The spread of the outbreak continues (4). According to WHO, the COVID-19 pandemic has resulted in 47,596,852 confirmed cases and 1,216,357 related deaths globally as of November 05, 2020; the number of cases and deaths continues to increase (2). Accordingly, the total number of cases and the number of deaths were 384,509 and 10,558, respectively, in Turkey (5).

Ambient and indoor particulate matter (PM) is a complex matrix that may contain various chemical and biological constituents (bacteria, virus, and fungi etc.) of a great health concern (6-15). Medium- and long-range transmission of bacteria and virus species on the atmospheric PM have been recently studied (11, 16-20). Accordingly, aerosol and droplets generated during speaking, sneezing, or coughing by infected people are well-known as the source of short-range transmission pathways for viral infections (21, 22). In particular, respiratory viral diseases can spread directly or indirectly through a virus-containing particle (droplet) among humans (23-26).

The primary transmission mode of COVID-19 is person-to-person contact through respiratory droplets generated by breathing, sneezing, coughing, and contact with an infected subject. Indirect contact with contaminated surfaces also transfers the virus to the mouth, nose, and eyes (27, 28). Transmission can also be through inhalation of the exhaled virus in respiratory droplets because of the long-term survival of coronaviruses outside of its host organism (29, 30).

Fiorillo et al. (31) reviewed several reports about the persistence of SARS-CoV-2 on various materials such as aerosols (3 hours), plastic (2 to 9 days), stainless steel (2 to 5 days), cardboard (8 to 24 hours), glass (4 to 5 days), and silicon rubber (5 days) and concluded that the persistence and transmission potency of the virus should not be underestimated. Buonanno et al. (32) estimated the quanta emission rates of SARS-CoV-2 emitted from contagious subjects based on activity patterns to implement airborne dispersion of the virus in indoor environments. According to their results, higher emission rates were achieved by an asymptomatic COVID-19 subject during both light and heavy exercise conditions as speaking or oral breathing whereas the symptomatic subject in resting conditions mostly has low emission rates. Similarly, Liu et al. (1) investigated the aerodynamic nature of SARS-CoV-2 in size-segregated indoor aerosol samples collected from different divisions of hospitals in Wuhan, China. They found very low SARS-CoV-2 RNA concentrations in isolation wards and aerated patient rooms with higher concentrations in patients’ toilets. They also reported that the size distribution of the SARS-CoV-2 peaked on the smallest particles suggesting that the long-range transmission was a possible spreading route due to the longer atmospheric lifetime of submicron aerosols (32-34). Finally, Prather et al. (35) remarked on the growing evidence of airborne transmission of SARS-CoV-2 as a major route of exposure among people.

Aerosolized respiratory viruses have great potential for spreading the infections (36-40). Several studies suggest that the spread of the infection may be related to the ambient air pollutant concentrations; such pollution could be a possible virus carrier. Correspondingly, COVID-19 may have a contagion route via airborne transmission on atmospheric PM (41-43). In a novel study, Setti et al. (44) obtained 20 positive results of the marker genes of SARS-CoV-2 among 34 ambient PM10 samples in the Bergamo area of Northern Italy suggesting a potential indicator of the transmission of the infection by ambient particles. However, there is still little known about the aerosol transmission of COVID-19 and the presence of virus on PM outdoors. The mode of transmission of the infection must be precisely determined to curtail the pandemic. In this study, we collected ambient PM samples in various size ranges from 13 sites in ten Turkish cities including urban and suburban locations as well as hospital gardens from May 13 to June 14, 2020. We demonstrated the presence of SARS-CoV-2 on PM collected from the various locations studied.

## Results

### Presence of the Virus on Ambient Atmospheric Particulate Matter

A total of 155 samples including TSP (*n = 80*), PM_2.5_ (*n = 33*), PM_2.5-10_ (*n = 23*), and PM_10_ (*n = 19*) were collected. Additionally, 48 PM samples segregated in 6 sizes were collected during the 8 days of sampling in urban İstanbul. **Table 1** shows the positive results of the RdRP and N1 genes on filters from various sites inspected within this study. The virus presence together with environmental parameters of all samples collected at each location are given in **Table S1** in *Supplementary Information (SI)*.

**Table 1.** RdRP and N1 gene positivity of filter samples.

| City | Site | Date | PM Size | RdRP gene (RT-PCR – Ct) | N1 gene (RT-PCR – Ct) | N1 gene (3D PCR – copy $\mu\text{L}^{-1}$) | Copy number on filter |
| --- | --- | --- | --- | --- | --- | --- | --- |
| Zonguldak | HG | 21.05.2020 | PM<sub>2.5</sub> | 35.99 | 44.14 | - |  |
| Zonguldak | HG | 23.05.2020 | PM<sub>2.5</sub> | 35.85 | 42.50 | - |  |
| Zonguldak | HG | 25.05.2020 | PM<sub>2.5</sub> | 35.90 | 42.21 | 14.615 | **118** |
| Zonguldak | HG | 26.05.2020 | PM<sub>2.5</sub> | 34.87 | 41.63 | - |  |
| Zonguldak | HG | 27.05.2020 | PM<sub>2.5</sub> | 35.34 | 42.91 | 30.655 | **246** |
| Zonguldak | HG | 27.05.2020 | PM<sub>10</sub> | 35.68 | 41.44 | - |  |
| Zonguldak | HG | 28.05.2020 | PM<sub>2.5</sub> | 35.57 | >45 | 19.773 | **158** |
| Zonguldak | HG | 28.05.2020 | PM<sub>2.5-10</sub> | 35.36 | >45 | 22.991 | **184** |
| Tekirdağ | HG | 17.05.2020 | PM<sub>2.5</sub> | 35.89 | 41.93 | - |  |
| Tekirdağ | HG | 18.05.2020 | PM<sub>2.5</sub> | 36.03 | 42.77 | - |  |
| Tekirdağ | HG | 19.05.2020 | PM<sub>2.5</sub> | 34.50 | 42.75 | 23.342 | **186** |
| Tekirdağ | HG | 20.05.2020 | PM<sub>2.5</sub> | 34.48 | 43.49 | 38.981 | **312** |
| Tekirdağ | HG | 21.05.2020 | PM<sub>2.5</sub> | 34.84 | 41.20 | 17.387 | **140** |
| Tekirdağ | HG | 27.05.2020 | PM<sub>2.5</sub> | 35.43 | 43.91 | 16.950 | **136** |
| Tekirdağ | HG | 28.05.2020 | PM<sub>2.5</sub> | 35.04 | 42.79 | - |  |
| Tekirdağ | HG | 29.05.2020 | PM<sub>2.5</sub> | 35.82 | 43.63 | 63.060 | **504** |
| Tekirdağ | HG | 30.05.2020 | PM<sub>2.5</sub> | 34.77 | 43.14 | 30.221 | **242** |
| Tekirdağ | HG | 31.05.2020 | PM<sub>2.5</sub> | 35.65 | 43.40 | - |  |
| Eskişehir | U | 30.05.2020 | TSP | 40.00 | 43.33 | 19.097 | **152** |
| İstanbul | U | 14.05.2020 | PM<sub>>7.2</sub> | **38.34** | **36.16** | 15.797 | **126** |
| İstanbul | U | 15.05.2020 | PM<sub><0.49</sub> | 37.32 | >45 | 43.790 | **350** |
| İstanbul | U | 15.05.2020 | PM<sub>>7.2</sub> | **37.37** | **37.49** | 9.917 | **80** |
| İstanbul | U | 20.05.2020 | PM<sub>0.49-0.95</sub> | 36.93 | >45 | - |  |
| İstanbul | U | 20.05.2020 | PM<sub>1.5-3</sub> | 37.19 | >45 | - |  |
| İstanbul | U | 27.05.2020 | PM<sub><0.49</sub> | 38.52 | >45 | - |  |
| İstanbul | U | 28.05.2020 | PM<sub>0.49-0.95</sub> | **38.01** | **35.29** | 12.056 | **96** |
| İstanbul | U | 28.05.2020 | PM<sub>0.95-1.5</sub> | **38.66** | **35.09** | 21.499 | **172** |
| İstanbul | U | 02.06.2020 | PM<sub><0.49</sub> | 37.15 | >45 | - |  |
| İstanbul | U | 02.06.2020 | PM<sub>0.49-0.95</sub> | 37.24 | >45 | - |  |
| İstanbul | U | 02.06.2020 | PM<sub>0.95-1.5</sub> | 36.92 | 44.41 | - |  |
| İstanbul | U | 02.06.2020 | PM<sub>3-7.2</sub> | 38.93 | 44.70 | - |  |
| İstanbul | HG | 13.05.2020 | PM<sub>2.5</sub> | 36.64 | >45 | 12.943 | **102** |
| İstanbul | HG | 14.05.2020 | TSP | 35.17 | >45 | - |  |
| İstanbul | HG | 14.05.2020 | PM<sub>2.5</sub> | 35.02 | >45 | - |  |
| İstanbul | HG | 20.05.2020 | TSP | 35.79 | >45 | - |  |
| İstanbul | HG | 20.05.2020 | PM<sub>2.5</sub> | 34.74 | 44.54 | - |  |
| İstanbul | HG | 21.05.2020 | TSP | 36.420 | >45 | 15.846 | **126** |
| İstanbul | HG | 02.06.2020 | TSP | >45 | >45 | 15.800 | **126** |
| Ankara | U | 30.05.2020 | TSP | >45 | >45 | 14.747 | **118** |

HG, Hospital garden; U, Urban site

The positivity of the SARS-CoV-2 on aerosol samples was characterized by a multi-parameter decision approach. **Figure 1** demonstrates the protocol for analysis of PM samples. Initially, the N1 gene was studied on all 203 PM samples including segregated ones by RT-PCR using Syber Green method. Of these, four samples were positive for the N1 gene with a Ct < 40 whereas 199 samples had a Ct value of ≥40 for the same gene. The latter were further analyzed for specific products by checking melting curves as well as specific products in a 2% agarose gel. Traces of specific products were detected in 52 samples, and these were further analyzed for RdRP genes together with four samples positive for the N1 gene by RT-PCR using the Taqman hybridization probe. Of these 56 samples, 4 samples had a Ct <40 for both N1 and RdRP genes, 33 samples had a Ct of ≥ 40 for N1 and Ct <40 for RdRP genes, and two samples had a Ct≥40 for both N1 and RdRP genes with a specific product in the gel for N1 gene. In total, 39 out of 56 samples were analyzed by 3D-dPCR for N1 gene, and 20 samples were positive for the N1 gene. The remaining 17 of 56 samples had a Ct≥40 for both N1 and RdRP genes with no sign of a specific product, and these samples were treated as negative for SARS-CoV-2. These 20 samples were amplified above 10 copies µL^-1^ and considered positive. Among the positive samples, the lowest copy number was 80 while the highest copy number was 504 copies on the filters (Table 1)

**Figure 1.**
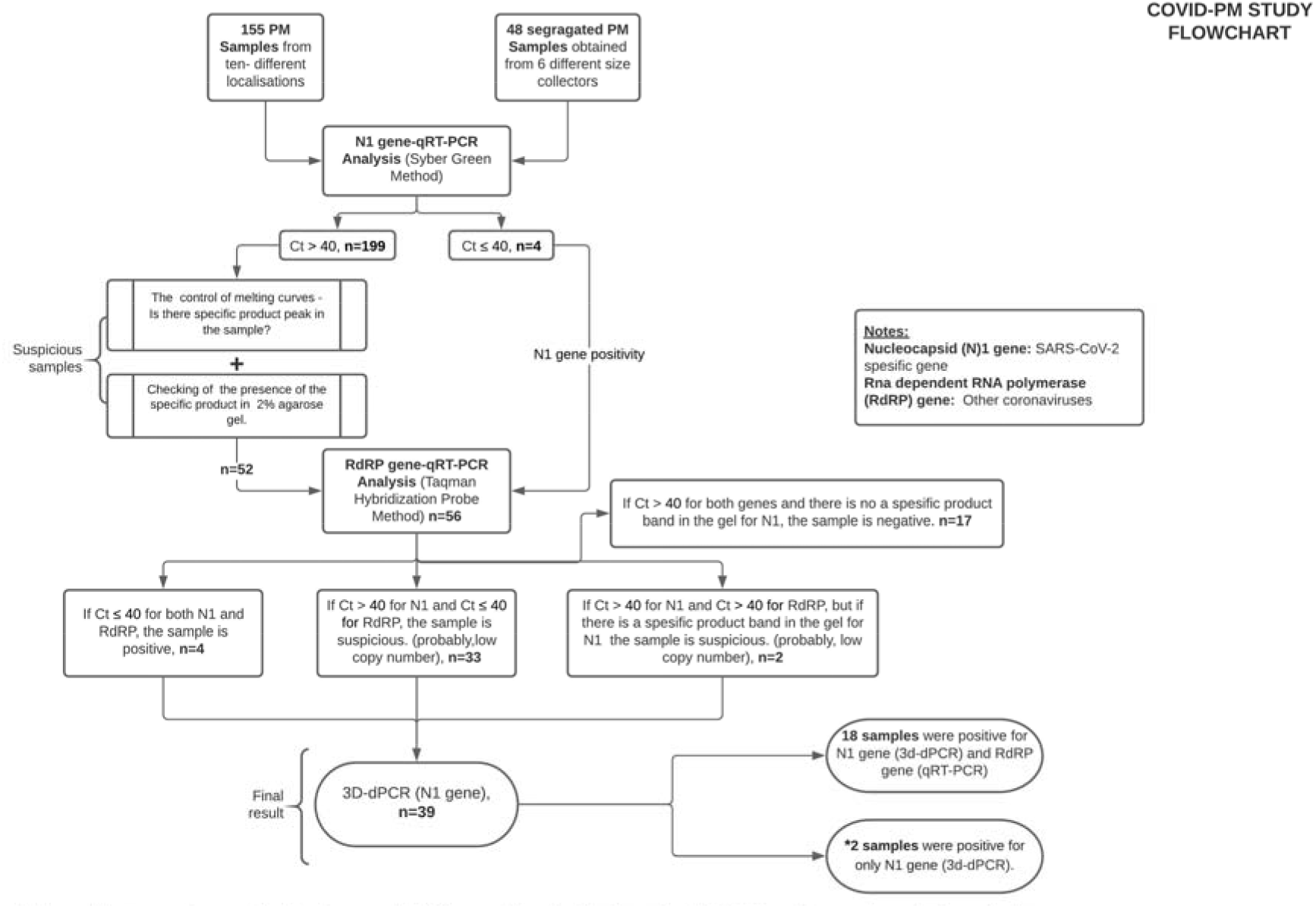
The flow-chart for analysis of particulate matter samples for N1 and RdRP genes.

Forty-eight PM samples were collected by an impactor having different size ranges (0.49 to >7.2 µm [0.49-0.95; 0.95-1.5; 1.5-3; 3-7.2 and >7.2 µm and a back-up filter <0.49 µm]) in urban İstanbul and analyzed for the virus. Among these size-distributed samples, the virus was detected only on five filters. The PM sizes of positive samples and the number of samples detected in size-segregated samples were PM_<0.49_ (*n = 1*), PM_0.49-0.95_ (*n = 1*), PM_0.95-1.5_ (*n = 1*), and PM_>7.2_ (*n = 2*). The virus was detected in size-segregated PM samples collected on three different days out of 8. On two succeeding days, the virus was detected on particles >7.2 µm and on <0.49 µm and >7.2 µm for 14^th^ and 15^th^ of May, respectively. Due to the lower percentage and the random results of positive observations in size-segregated samples, we could not observe a systematic distribution according to PM sizes. Thus, a detailed description of the size distribution of SARS-CoV-2 in atmospheric particles needs more attention and systematic approaches for further research. When background samples were analyzed neither the N1 nor the RdRP gene was amplified during the 45 cycles of QRT-PCR. Therefore, these samples were negative for the presence of viruses, and no interference was observed from the used filters and equipment. In the rest of 155 PM samples, positive counts of SARS-CoV-2 RNA on other ambient particles according to PM sizes were: PM_2.5_ (*n = 10*), PM_10_ (*n = 1*), and TSP (*n = 4*) (**Table 1**).

Of the samples positive for SARS-CoV-2, 13 samples were close to hospitals, and the remaining 7 positive results were from urban sites. The locations of positive samples included Zonguldak (*n = 4*, hospital garden), Tekirdağ-Çorlu (*n = 6*, hospital garden), İstanbul site 1 (*n = 3*, hospital garden), Eskişehir (*n = 1*, urban), İstanbul (*n = 5*, urban), and Ankara (*n = 1*, urban). All analyzed samples were negative for the virus collected from Bolu (urban and urban background), Bursa (urban background and hospital garden), Konya (hospital garden), Antalya (urban background), and İzmir (urban site).

## Discussion

We demonstrated that SARS-CoV-2 RNA can be present on ambient PM suggesting that this virus may be transported via PM pollution. **Figure 2** shows the ambient air particle-bound SARS-CoV-2 concentrations for sampling sites excluding the size segregated samples of the urban site of İstanbul in terms of N1 gene copy number (copy m^-3^) calculated using the sampling volumes in **Table S1** for each sample. It is clear from **Table 1** and **Figure 2** that positive results were obtained from the hospital gardens for most of the samples (60%, 50%, and 30% of total samples in Tekirdağ, İstanbul, and Zonguldak, respectively, and especially in PM_2.5_ mode). However, most of the SARS-CoV-2-positive samples were collected during the lockdown time. During the sampling period, there was a lockdown precaution in 9 of the 10 sampling cities due to increasing daily COVID-19 cases. Daily cases and cases per capita were also high in those 9 cities especially in İstanbul, Ankara, İzmir, and Zonguldak exceeding national average. Although the population and daily COVID-19 cases were higher in İzmir and Bursa cities (populations of 4,321,000 and 2,995,000, respectively), there were no positive samples detected. Among the sampling locations, Tekirdağ and Zonguldak (populations of 1,030,000 and 213,544, respectively) are smaller cities compared to others but have worse air pollution (45). Tekirdağ is an industrialized city and Zonguldak is a coal mining area with coal-fired power plants.

**Figure 2.**
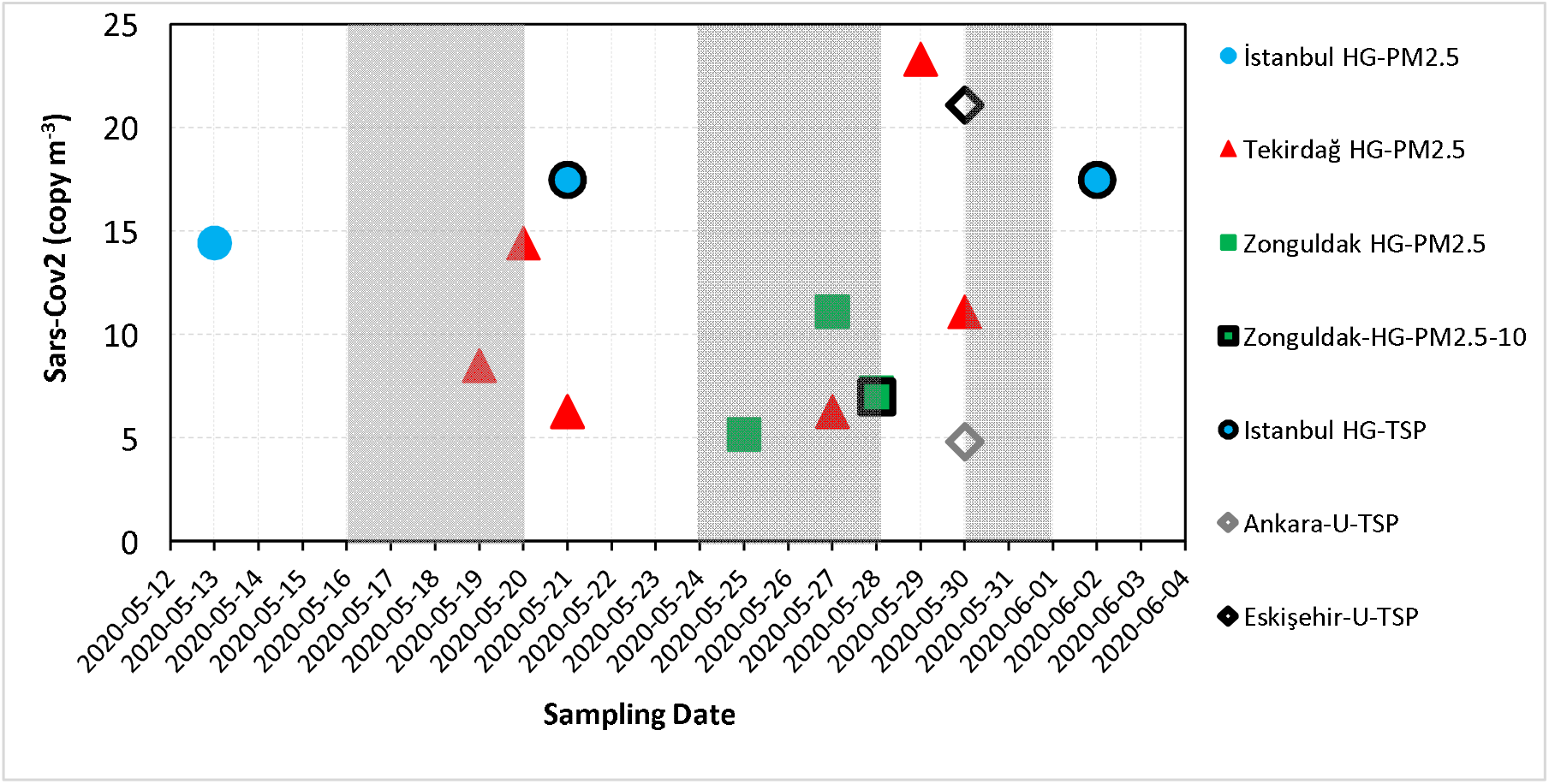
SARS-CoV-2 copy number on TSP, PM_10_, PM_2.5-10_ and PM_2.5_ for sampling sites (excluding size segregated samples). The grey shaded area on the graph shows the lockdown periods in Turkey. The indicator colors of blue, red, green, black and grey represents the samples from İstanbul, Tekirdağ, Zonguldak, Ankara and Eskişehir, respectively. HG, hospital garden, U, urban sites; TSP, total suspended particulate; PM, particulate matter.

Airborne transmission of SARS-CoV-2 was previously investigated by Bontempi (41) in the ambient atmosphere of Northern Italy in relation to the increasing concentrations of PM. After a basic correlation analysis of ambient PM data and confirmed cases obtained from the cities of Brescia, Bergamo, Cremona, Lodi, Milano, Monza-Brianza, Pavia, Alessandria, Vercelli, Novara, Biella, Asti, and Torino, they reported that it was not possible to conclude that the infection was transmitted through PM10 particles without any additional information. Consequently, Setti et al. (46) reported the presence of the SARS-CoV-2 RNA on ambient aerosol samples collected from the ambient air of Bergamo (Northern Italy) where the incidence of the COVID-19 was extremely high during February 21^st^ and March 13^th^, 2020. In contrast to Bontempi’s study, they inspected the markers of the virus on ambient particles rather than the PM mass concentrations alone. They reported 20 positive results for at least one marker among the 34 RNA extractions for three marker genes (E, N, and RdRP).

There are also several reference works on the transmission of influenza viruses by PM (10, 11, 16 - 18, 20, 36). Bao et al. (47) investigated the transmission of influenza viruses by atmospheric particles using detailed surface characterization of ambient fine particles and an advanced transmission technique (synchrotron-based transmission X-ray microscope). They proposed that the long-range transport of influenza viruses is possible by combustion derived particles, which have pores providing a suitable space for the virus settlement. This strong assumption may also be valid for our case since a high incidence of daily COVID-19 were observed in Zonguldak and Tekirdağ cities where extreme coal and industrial particles were reported (48-50) as well as in İstanbul—the biggest metropolitan city of Turkey suffering from traffic and industry-related air pollution (51, 52).

The presence of the SARS-CoV-2 RNA on aerosol samples in indoor environments has previously been studied by several researchers (53-59). It is well documented that the indoor concentration of the virus may reach significant levels especially in hospital rooms and corridors; therefore, this is an important route of exposure for health professionals working in intensive care units. Droplets generated during oral activities like breathing, speaking, singing, coughing, sneezing, and clinical dental practices are known to be the major carriers of the virus (28, 32, 53-56). Correspondingly, two indoor super spreading cases due to the inhalation of SARS-CoV-2 aerosol were reported from a call center in South Korea (57) and Skagit Valley Chorale, Washington, USA (60). Of 216 employees of the call center, 94 tested positives for COVID-19; 53 of 61 members of the Skagit Valley Chorale were infected.

The amount of suspended viral copies in the air is another significant factor affecting the airborne transmission of SARS-CoV-2. Lednicky et al. (59) reported indoor air virus presence from a hospital room with two COVID-19 patients. They reported 16 to 94 SARS-CoV-2 genome equivalents per L of indoor air in the patients’ room after 3-h indoor PM samplings. Liu et al. (1) investigated the aerodynamic nature of the virus in two Wuhan hospitals from February to March 2020 by collecting indoor aerosol samples with a cascade sampler and a similar virus detection protocol as in the present study. They reported indoor air virus concentrations of 0 to 11 copies per m^3^ on the PM samples collected from public areas, 0 to 42 copies per m^3^ on medical staff areas, and 0 to 113 copies per m^3^ in the patient areas including the toilets, workstations, and intensive care units.

In a similar study, both indoor and ambient air samples were collected and analyzed for the presence of SARS-CoV-2 from selected microenvironments in Wuhan, China (60). Positive viral RNA percentages were 10% and 20% for the ambient samples collected from a 10-m distance to the inpatient and outpatient building doors, and the air concentrations were 0.89 to 1.65 × 10^3^ copies m^-3^. Those results support the findings of the present study regarding the higher percentage of detection of the virus on PM samples from hospital gardens in İstanbul, Tekirdağ, and Zonguldak. The location of the PM samplers was on close proximities to the hospital main buildings in most of the sampling cities except Bolu (urban and urban background), Antalya (sub-urban), Eskişehir (urban), İstanbul site 1 (urban), Ankara (urban), and İzmir (urban). The sampler in Tekirdağ was a close spot to the exhaust fans of the main ventilation system of the building and showed the highest positive virus incidences. According to the results of this study and in the light of the previous studies reporting virus prevalence, locations close to the hospitals in populated urban areas may be the significant zones for the airborne transmission of SARS-CoV-2 viruses. Due to the higher occurrence of the positive virus, RNA copies—especially on fine mode particles—are also asserting the idea that the airborne transmission of the SARS-CoV-2 through respirable particles is possible if the environmental parameters (PM surface characteristics, temperature, sunlight intensity, and humidity) are favorable for the viability of the virus. Although the air concentration of the SARS-CoV-2 RNA in ambient air was comparably lower than those reported in the studies by Hu et al. (60) and Liu et al. (1), various concentrations between 5 RNA copy m^-3^ (Ankara, TSP) and 23 RNA copy m^-3^ (Tekirdağ, PM2.5) were observed in the present study suggesting that transmission of the virus through the aerosols should not be ignored even in the ambient air.

Basic air quality parameters like PM, sulfur dioxide (SO_2_), nitrogen oxides (NO_x_), ozone (O_3_), carbon monoxide (CO), volatile organic compounds (VOCs), and polycyclic aromatic hydrocarbons (PAHs) together with meteorological parameters were all inspected to have a potential effect on the transmission of respiratory infections in the literature. Domingo and Rovira (20) reviewed the scientific literature on ambient air pollution and respiratory diseases. The review supported an obvious relationship between ambient levels of certain pollutants and viruses influencing each other to negatively deteriorate the human respiratory system. Liu et al. (61) compiled a daily confirmed case count, ambient temperature, diurnal temperature range, absolute humidity, and migration scale index data from 30 provincial capital cities of China to explain the associations between COVID-19 and meteorological parameters using a non-linear regression analysis. They concluded that the meteorological parameters play an independent functionality in the COVID-19 transmission after controlling for population migration. Local meteorological conditions with low to mild diurnal temperatures together with low humidity likely favored the transmission.

In a similar study, Zhang et al. (43) inspected ambient meteorology and local air quality on the distribution of the disease in 219 Chinese cities from January 24^th^ to February 29^th^, 2020. They similarly concluded a negative relationship between ambient temperature and positive correlations among air pollution indicators and confirmed cases. Indeed, both studies were conducted on linear or non-linear statistical relationships with daily confirmed cases and environmental inputs, and presented stochastic results among model inputs. The results could not statistically be related to the ambient air quality or meteorological parameters. Even though the total number of filter samples collected (in total, 155 PM samples) was relatively high, the number of individual samples from each site varied between 7 and 20 and limited the construction of a statistical model for a correlation analysis. Because of the restrictions on disseminating specific data on COVID-19 cases, data from each individual city could not be obtained—only the number of total national cases were shared with the public. Therefore, a possible association between the presence of SARS-CoV-2 in PM samples from sites studied and the number of patients reported from those areas could not be investigated.

A summary of mean air quality and the meteorological parameters in the PM sampling cities are given in **Table S2**. Relatively lower pollutant concentrations were obtained due to the decreased anthropogenic activity during the pandemic precautions, stay-at-home advice, and lockdown measures from a nationwide perspective (62). A variable ambient temperature range was observed among 10 cities during PM sampling due to the transitional characteristics of the spring season. Daily average temperatures, wind speeds, relative humidity (RH) ranges, as well as precipitation levels all differed substantially, and all were affected by the basic air quality parameters especially PM concentrations. Besides, the synergistic combination of ambient meteorological parameters (temperature and RH), air quality (PM toxicity) may also have a significant effect on vitality and transmission of aerosol biological constituents characterized by the suitability and atmospheric lifetime/travel distance of suspended particles (20). Correspondingly, the RH values observed in İstanbul (76.3%), Zonguldak (75.0%), and Tekirdağ (72.4%) were all higher than those reported from remaining cities (varying between 51.9% and 67.2%). The results were not sufficient to analyze the impact of humid or PM levels on the transmission of the virus adsorbed on particles. Both positive and negative non-linear relationships between ambient temperatures and COVID-19 cases were reported in different cities in China by Zhang et al. (43). More importantly, they reported direct positive correlations between new confirmed cases and air pollution indicators as viral spread increased with air quality index (AQI).

This study offers a preliminary evaluation of the possible presence of SARS-CoV-2 on ambient PM in characteristic sites of Turkey during the most intense period of COVID-19 pandemic. Our findings demonstrated that ambient PM collected outdoors could contain SARS-CoV-2; however, the vitality of the detected virus could not be assessed within the scope of this study. Most of the SARS-CoV-2 positive samples were from hospital gardens whereas samples collected from two urban sides were also positive. Although various studies from the literature suggest a relation between meteorological parameters, air quality parameters, and SARS-CoV-2 transmission, the current study shows that the data on these parameters were not adequate to perform a statistical analysis. Nevertheless, our findings suggest that SARS-CoV-2 may be transported by ambient particles—whether this has an impact on the spread of the virus remains to be determined. Therefore, the public should use personal protection equipment such as face masks during outdoor activities. Future studies should focus on the viability and infectivity of the SARS-CoV-2 present on both indoor and ambient particles.

## Materials and Methods

### Particulate matter sampling locations and methods

PM samples within various size-ranges were collected from 13 locations within 10 cities in western Turkey between 13^th^ May and 14^th^ June 2020 during the first peak of the outbreak in Turkey. **Figure 3** shows the location map of PM sampling sites, and **Table 2** summarizes the methodology of PM sampling, cut sizes of collected PM, and the typical features of the sampling locations. A total of 155 samples (TSP, *n=80*; PM_2.5_, *n=33*; PM_2.5-10_, *n=23*; PM_10_, *n=19*) were collected daily using various PM samplers in each city. Furthermore, 24-h size-segregated PM samples (*8 days*) were collected by a six-stage impactor system in the urban areas of İstanbul (Esenler) to investigate the fractional differences of the possible presence of the SARS-CoV-2. The impactor system consists of 5 stages and a back-up stage for size cutting from 0.49 to >7.2 µm (0.49-0.95; 0.95-1.5; 1.5-3; 3-7.2 and >7.2 µm and a back-up filter <0.49 µm). Therefore, 6 samples of different size fractions per day were collected at this site. Five of the 13 sampling sites were in the hospital gardens. Five sites were characterized as urban, and the remaining three sites were urban background locations. Samples were collected on glass fiber filters (GF) and Teflon filters (TF) with different sampling equipment (**Table 2**). Both low and high-volume samplers were used to collect daily samples. PM samples were collected by research teams of several universities of Turkey and transported to, biosafety level 2 microbiology research laboratory at Koç University, School of Medicine for PCR analysis.

**Table 2.** The descriptions of particulate matter sampling methods from 13 sites in 10 cities in Turkey.

| Cities | Site | PM Sampling Equipment | Sampling period | Number of samples | PM Fraction | Filter Types | Sampling volume, m<sup>3</sup> day<sup>-1</sup> |
| --- | --- | --- | --- | --- | --- | --- | --- |
| İstanbul | HG | SKC Filter Pack Sampler | 13.05.2020-04.06.2020 | 20 | TSP and PM<sub>2.5</sub> | PTFE | 7.2 |
| Tekirdağ | HG | Dichotomous PM Sampler | 17.05.2020-31.05.2020 | 20 | PM<sub>2.5-10</sub> and PM<sub>2.5</sub> | PTFE | 24 |
| Bursa | HG | High Volume Air Sampler | 21.05.2020-27.05.2020 | 7 | TSP | GF | 278 |
| Zonguldak | HG | Dichotomous PM Sampler | 16.05.2020-28.05.2020 | 26 | PM<sub>2.5-10</sub> and PM<sub>2.5</sub> | PTFE | 24 |
| Konya | HG | High Volume Air Sampler | 21.05.2020-03.06.2020 | 14 | TSP | GF | 360 |
| Ankara | U | Low Vol Stack Filter Unit | 30.05.2020-14.06.2020 | 10 | TSP | PTFE | 24 |
| Bolu | U | SKC Filter Pack Sampler | 14.05.2020-22.05.2020 | 9 | TSP | PTFE | 7.2 |
| İzmir | U | Zambelli PM Sampler | 14.05.2020-02.06.2020 | 9 | PM<sub>10</sub> | PTFE | 24 |
| İstanbul | U | High Vol Cascade Impactor | 13.05.2020-02.06.2020 | 48 | 0.49-7.2 µm (6 fractions) | GF | 1422 |
| Eskişehir | U | SKC Filter Pack Sampler | 16.05.2020-30.05.2020 | 14 | TSP | PTFE | 7.2 |
| Bursa | UB | High Volume Air Sampler | 21.05.2020-27.05.2020 | 7 | TSP | GF | 318 |
| Bolu | UB | SKC Filter Pack Sampler | 14.05.2020-22.05.2020 | 9 | TSP | PTFE | 7.2 |
| Antalya | UB | Low Vol Stack Filter Unit | 18.05.2020-03.06.2020 | 10 | PM<sub>10</sub> | NPF | 24 |

U: Urban site, UB: Urban background, HG: Hospital garden, GF: Glass fiber filter, PTFE: Teflon filter, NPF: Nuclepore Polycarbonate filter, TSP: Total Suspended Particulate, PM<sub>2.5-10</sub>: Particulate Matter between 2.5 and 10 µm, PM<sub>10</sub>: Particulate Matter <10 µm.

**Figure 3.**
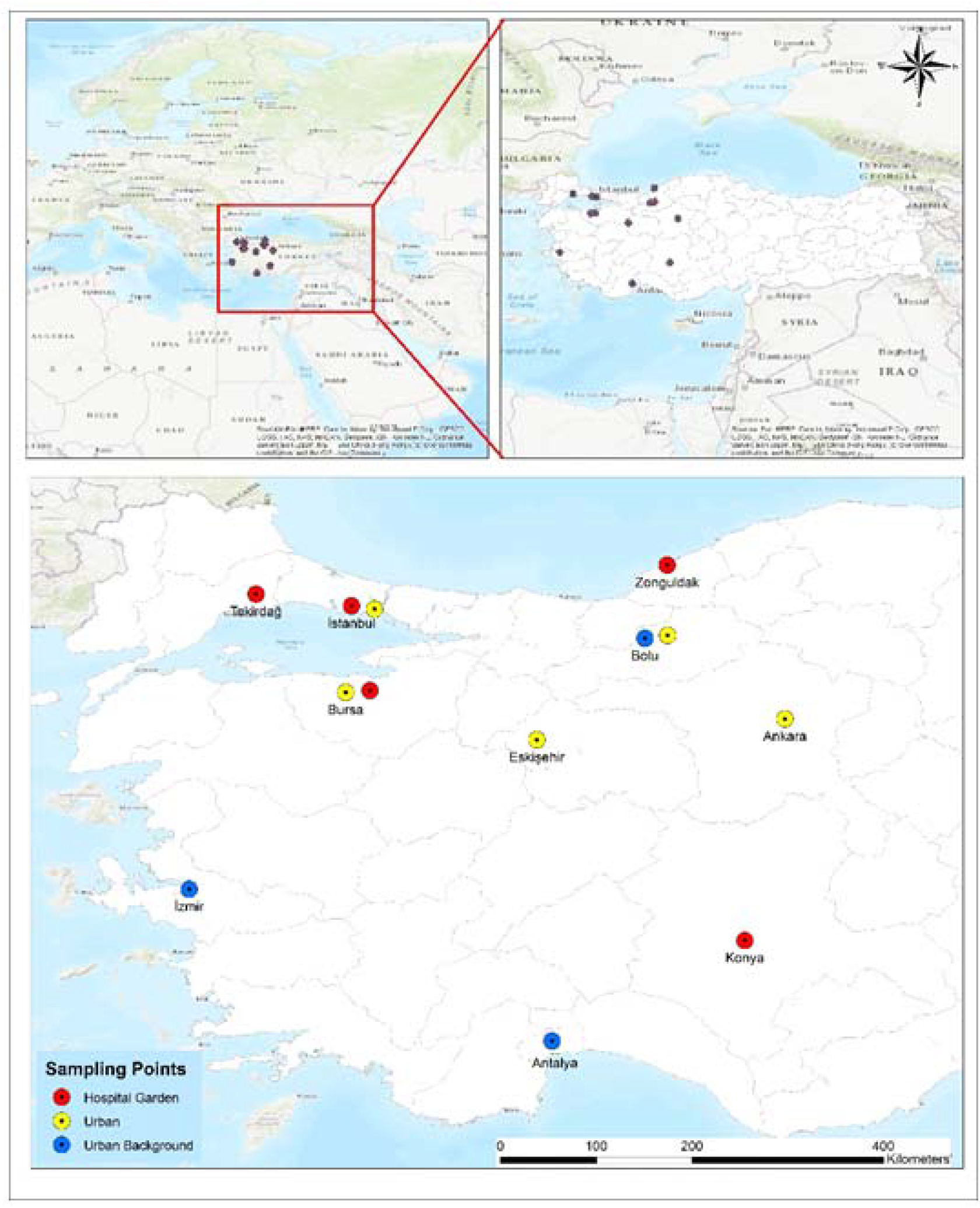
Sampling locations in Turkey.

For all sampling locations, sampling started in the morning and finished at the same time on the following day. At the end of 24 hrs, each sample was placed into sterile petri dish using sterile equipment and stored at −20° C. Filters were cut into halves with a sterile scalpel: One half was immediately subjected to PCR analysis. The remaining halves of the filters were kept frozen for further chemical and bacteriological characterization. Since the main purpose of this study was to investigate the presence of SARS-CoV-2, filters were not weighed to avoid any contamination. Field blanks (*n=3*) from each site were also analyzed together with samples to detect any contamination.

There are more than 300 air quality monitoring stations in Turkey operated by the Turkish Ministry of Environment and Urbanization (MoEU) according to European and United States standards. To evaluate the air quality and meteorology in the PM sampling cities during the PM sampling time, daily averages of PM_2.5_, PM_10_, NO_2_, SO_2_, CO, and O_3_ were acquired from the national air quality monitoring network of the MoEU (63), and the meteorological parameters were obtained from the National Oceanic and Atmospheric Administration (NOAA) using the locations of PM sampling sites (64).

### Analytical methods

#### RNA Isolation

RNA isolation was performed using the Quick-RNA™ Fecal/Soil Microbe Microprep Kit (ZYMO Research, USA) according to the manufacturer’s instructions. Briefly, the half filter was rolled with the upper surface facing inward in a 2-mL polypropylene tube together with the beads provided in the kit. From the initial 1 mL of lysis buffer, ∼400 µL of lysate was harvested, and then processed as defined by the instructions resulting in a final eluate of 10 µL. Subsequently, all the eluted RNA was used for SARS-CoV-2 testing.

#### cDNA synthesis and Quantitative Real-Time Polymerase Chain Reaction (QRT-PCR)

Quantitative real-time polymerase chain reaction (QRT-PCR) is a real-time method that can give absolute or relative results that detects the amount of DNA doubled in the previous cycle during each amplification process with a fluorescent detector such as Syber Green and FAM probe systems (65). cDNA was synthesized from the eluted RNA with VeritiTM Thermal Cycler (Applied Biosystem, USA) using iScript™ cDNA Synthesis Kit (Bio-Rad, USA) according to the instructions of the manufacturer. Five µL of cDNA was used for qRT-PCR analyses. Given the scarcity of cDNA, we decided to test for the presence of SARS-CoV-2 RNA on airborne particles with two different specific markers (nucleocapsid [N]1 and RNA dependent RNA polymerase [RdRP]) genes (66, 67). Samples with threshold cycle (Ct) values of 40 and below for the N1 and RdRP genes were identified as positive. Samples with Ct values of 40 and above for RdRP and/or N1 gene were identified as suspicious samples. First, the N1 gene was tested using Syber Green Master Mix II (Roche, Germany) in all collected samples on the LightCycler 480 Real-Time PCR system (Roche, Germany). The N1 gene cycle of the Ct value ≤ 40 was considered positive. Then RdRP gene expressions were analyzed in the N1 positive and/or suspicious samples using Taqman Hybridization Probe Master Mix (Roche, Germany) on the same QRT-PCR system. The samples that have dual positivity with both genes were considered as SARS-CoV-2 positive. Cycling conditions for Syber detection were as follows: in 5 min at 95° C, followed by 45 cycles of 10 sec at 95° C, 30 sec at 63° C, and 30 sec at 72°C, and final cooling step at 4° C. Cycling conditions for Taqman Hybridization was as follows: in 7 min at 95° C, followed by 45 cycles of 10 sec at 95° C, 1 min at 63° C, and 30 sec at 72° C followed by maintenance at 4° C. Gene sets were obtained from the Centers for Disease Control and Prevention (CDC). Primer sequences: N1 Forward: 5’-GACCCCAAAATCAGCGAAAT-3’, N1 Reverse: 5’-TCTGGTTACTGCCAGTTGAATCTG-3’; as an internal control RdRP Forward:5’-AGATTTGGACCTGCGAGCG-3’, RdRP Reverse: 5’-GAG CGG CTG TCT CCA CAA GT-3’. Probe: FAM – 5’-TTC TGA CCT GAA GGC TCT GCG CG −3’– BHQ1 (66).

#### Three Dimensional (3D)-Digital PCR

The main difference of three dimensional (3D)-digital PCR (3D-PCR) from QRT-PCR is that the reaction volume is split over a high number of small partitions (from 500 up to millions) of a very small volume (currently from 6 nanoliters down to a few picolitres). After the PCR, each partition is scored either as positive or negative (binary or digital read-out). Statistical analysis of the results is then used to determine the absolute quantity of target DNA in a sample (68). This method shows high sensitivity with the strategy of counting a single molecule. It also provides a high reliability and repeatability level. Furthermore, this method has been proven to work with high sensitivity in studies related to SARS-COV-2 (69).

Airborne PM may have an unknown interaction level with the virus resulting in a low or high copy number of the virus. Thus, we aimed to detect this interaction with highly sensitive-dPCR QuantStudio 3D™ System (Thermo, USA) in samples previously determined as positive or suspicious for the presence of RdRP and/or N1 genes (62, 69). Therefore, we further analyzed these samples for the existence of the N1 gene, which is specific to SARS-CoV-2 using QuantStudio™ 3D Digital PCR Master Mix v2 and QuantStudio™ 3D 20K v2 chips according to the instructions of the manufacturer (Thermo, USA).

Here, 5 µL of cDNA was used for 3D-dPCR analyses. Gene and probe sets for N1 gene were obtained from the CDC as described above (66). Primers used in real-time PCR analyses were also used in 3D-dPCR analyses. The probe sequence of N1 gene was FAM-5’-ACCCCGCATTACGTTTGGTGGACC-3’-BHQ1. Cycling conditions were as follows: in 10 min at 96° C, followed by 39 cycles of 30 sec at 63° C and 2 min at 98° C, and a final step of 63° C per 2 min followed by maintenance at 4° C. The chips were read in the QuantStudio 3D™ reader (Thermo, USA), and the results were interpreted in the dPCR AnalysisSuite™ app in the Thermo Fisher Connect™ Dashboard. Results with precision values ≤ 20% were selected to estimate the quantity of SARS-CoV-2 genomic copies based on 3D-dPCR (62).

## Data Availability

All data referred to in the manuscript can be available citing the manuscript.

## Acknowledgments

The authors thank Koc University Research Center for Translational Medicine (KUTTAM) for funding the laboratory analysis of SARS-CoV-2 in PM samples. The authors thank to officials of Zonguldak Bülent Ecevit University Science and Technology Research Center, and Health Medical Research and Application Center for device support and workforce support during the sampling studies.

